# Reliable Monitoring of Respiratory Function with Home Spirometry in People Living with Amyotrophic Lateral Sclerosis

**DOI:** 10.1101/2025.02.03.25321333

**Authors:** Julián Peller, Marcos A Trevisan, Gaston Bujia, Felipe Aguirre, Diego E. Shalom, Alan Taitz, Stephanie Henze, Silviya Bastola, Jason Osik, Ryan A. Shewcraft, Peng Jiang, Joel Schwartz, Terry Heiman-Patterson, Michael E. Sherman, Matthew F. Wipperman, Oren Levy, Guofa Shou, Karl A. Sillay, Lyle W. Ostrow, Ernest Fraenkel, James D. Berry, Indu Navar Bingham, Esteban G. Roitberg

**Author notes:** First coauthors (Julián Peller, Marcos A Trevisan).

## Abstract

Monitoring respiratory function is essential for assessing the progression of Amyotrophic Lateral Sclerosis (ALS) and planning interventions. Using spirometry data from the Radcliff Study —a fully remote, longitudinal, exploratory study with a cohort of 67 pALS—we demonstrate that flexible coaching, combined with a quality control analysis that excludes values of ‘0’ and timepoints with failed measurement trials, produces consistent remote spirometry results. Our findings indicate that home-measured Slow Vital Capacity (SVC) and Forced Vital Capacity (FVC) evolve similarly and progress linearly over the study period (7.7 ± 4.0 months). This remains true in both slow and fast progressor subpopulations. This observed linearity in respiratory trajectories supports the potential for early, accurate estimation of progression, reinforcing the feasibility of less frequent monitoring without compromising assessment precision, and reducing the burden on both pALS and the healthcare system. Furthermore, our results align with reported in-clinic pulmonary tests, validating remote monitoring as a means to promote more equitable and accessible clinical trial designs.

## Introduction

Amyotrophic Lateral Sclerosis (ALS) is a progressive neurodegenerative disease that affects nerve cells in the brain and spinal cord (Kiernan et al., 2011). As the disease advances, people living with ALS (pALS) experience muscle weakness and atrophy, ultimately leading to diffuse extremity weakness, bulbar difficulties and respiratory compromise. Monitoring respiratory function in pALS is crucial for assessing disease progression and planning interventions, such as non-invasive ventilatory support (Pinto and Carvalho, 2014). In the United States, approximately half of pALS receive care in multidisciplinary clinics where vital capacity and other pulmonary function tests are regularly conducted (Stegmann et al., 2021).

In-clinic interventions can be challenging due to several factors including the burden of transportation, which can be especially difficult for pALS in the advanced stages of the disease (Lin et al., 2023). Furthermore, many pALS live far from multidisciplinary clinics and trial centers, limiting their access to specialized care and research opportunities (Horton et al., 2018). Validating remote monitoring could significantly improve care delivery, facilitate research and promote equity in clinical trials.

The COVID-19 pandemic accelerated the transition to telemedicine, providing an appealing alternative to in-clinic visits by reducing participant burden and enabling more frequent evaluations (De Marchi et al., 2020). Studies comparing remote pulmonary function testing (rPFT) with in-clinic assessments indicate that rPFT requires optimized training and coaching for accurate measurements at home (Anand et al., 2023). Without ongoing reinforcement of home spirometry technique after initial training, discrepancies with clinic spirometry arise. Paynter et al. (2022) emphasize that the impact of coaching, setting, and equipment must be well understood to enhance home spirometry for conditions such as Cystic Fibrosis.

Research on rPFTs in patients with conditions such as motor neuron disease and chronic respiratory disease suggests that these tests are both accurate and acceptable when participants receive coaching, potentially facilitating the integration of home spirometries into telemedicine for clinical and research purposes (Geronimo and Simmons, 2019). Notably, remote monitoring provided almost two months’ advance notice for the need for non-invasive ventilation, compared to standard quarterly in-clinic assessments of respiratory function in pALS (Geronimo and Simmons, 2024). Wilson et al. (2024) demonstrated that home spirometry is an accurate and feasible method for assessing lung function, showing good agreement with desktop spirometry. Furthermore, unsupervised home spirometry following face-to-face training has been shown to be a valid and time-efficient method for remotely monitoring respiratory function, and is well-accepted by individuals and their caregivers (Helleman et al., 2022).

Coaching appears to be a key factor for obtaining accurate rPFTs, raising the question of what type of proctoring ensures valid home spirometry measurements. We address this in Radcliff Study (2024), a fully remote, observational, exploratory, and non-interventional study conducted at home by 67 participants. The Radcliff Study was designed to allow pALS to manage their coaching autonomously after an initial training period, enabling them to perform spirometry independently or request assistance from trained personnel. In this work, we show that combining this flexible coaching approach with a predefined automatic quality control protocol effectively ensures consistent and reliable spirometry results for studying respiratory function over time.

This work focuses on two pulmonary tasks: Slow Vital Capacity (SVC), which measures the total air volume that can be slowly exhaled after a deep breath, and Forced Vital Capacity (FVC), which measures the total air volume that can be forcibly exhaled during a rapid exhalation (Cotes et al., 2009). FVC is the most commonly used measurement to assess lung vital capacity (VC) (Czaplinski et al., 2006). In contrast, SVC does not require a minimum airflow, making it a potentially more accurate measure of VC in individuals with conditions that cause muscle spasticity, such as ALS (Pinto and de Carvalho, 2019; Huang et al., 2022).

## Results

Sixty-seven pALS participated in at least one session of the Radcliff Study. As the study required participants to independently perform respiratory, speech, and gait tasks, most participants were in the early stages of the disease. The mean age was 62 years (range: 51–68), with 57% male and 37% female participants (6% unspecified). The cohort was 90% white, a proportion higher than that reported in the US National ALS Registry, but lower than that reported in other US-based ALS trials (Cudkowicz et al., 2013) (see Table 1 for detailed demographic information). Each session included an initial set of respiratory tasks, followed by speech tasks, gait and balance assessments, a post-exercise speech evaluation, and a concluding set of post-exercise respiratory exercises (see Methods).

**Table 1:**
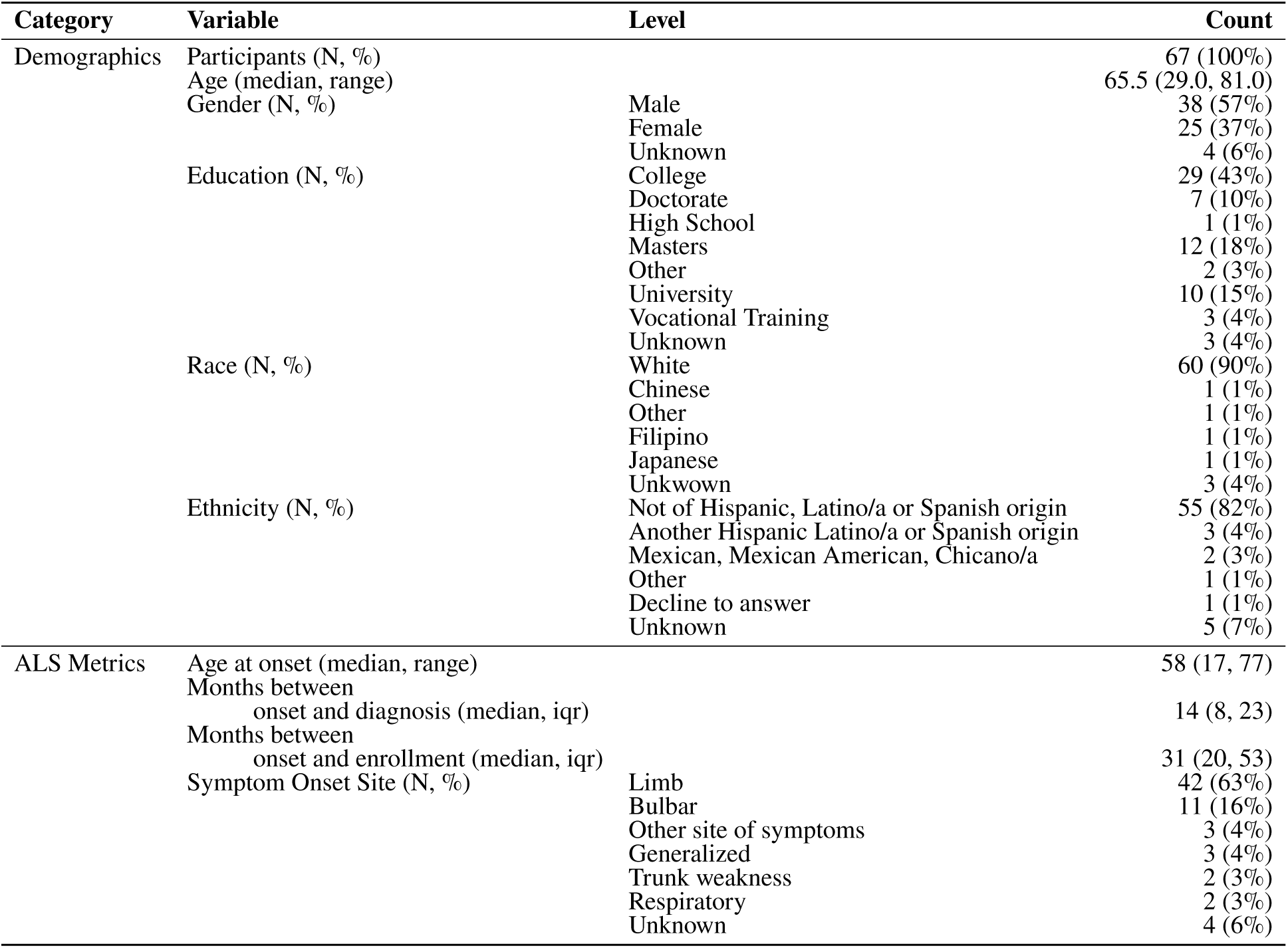
Table of Demographic and ALS-specific metrics.

| Category | Variable | Level | Count |
| --- | --- | --- | --- |
| Demographics | Participants (N, %) |  | 67 (100%) |
|  | Age (median, range) |  | 65.5 (29.0, 81.0) |
|  | Gender (N, %) | Male | 38 (57%) |
|  |  | Female | 25 (37%) |
|  |  | Unknown | 4 (6%) |
|  | Education (N, %) | College | 29 (43%) |
|  |  | Doctorate | 7 (10%) |
|  |  | High School | 1 (1%) |
|  |  | Masters | 12 (18%) |
|  |  | Other | 2 (3%) |
|  |  | University | 10 (15%) |
|  |  | Vocational Training | 3 (4%) |
|  |  | Unknown | 3 (4%) |
|  | Race (N, %) | White | 60 (90%) |
|  |  | Chinese | 1 (1%) |
|  |  | Other | 1 (1%) |
|  |  | Filipino | 1 (1%) |
|  |  | Japanese | 1 (1%) |
|  |  | Unkwown | 3 (4%) |
|  | Ethnicity (N, %) | Not of Hispanic, Latino/a or Spanish origin | 55 (82%) |
|  |  | Another Hispanic Latino/a or Spanish origin | 3 (4%) |
|  |  | Mexican, Mexican American, Chicano/a | 2 (3%) |
|  |  | Other | 1 (1%) |
|  |  | Decline to answer | 1 (1%) |
|  |  | Unknown | 5 (7%) |
| ALS Metrics | Age at onset (median, range) |  | 58 (17, 77) |
|  | Months between onset and diagnosis (median, iqr) |  | 14 (8, 23) |
|  | Months between onset and enrollment (median, iqr) |  | 31 (20, 53) |
|  | Symptom Onset Site (N, %) | Limb | 42 (63%) |
|  |  | Bulbar | 11 (16%) |
|  |  | Other site of symptoms | 3 (4%) |
|  |  | Generalized | 3 (4%) |
|  |  | Trunk weakness | 2 (3%) |
|  |  | Respiratory | 2 (3%) |
|  |  | Unknown | 4 (6%) |

Here we investigated the evolution of the respiratory outcome measures, focusing on the three efforts of SVC and FVC performed at the beginning of each session. To characterize the attendance to the study, we analyzed the subpopulation of 51 pALS who completed at least three sessions, which allows computing an average session frequency for each participant. The distribution of participants based on their permanence in the study and average session frequency is shown in Figure 1a, showing that most of the participants attended between 2 and 4 sessions per month.

**Figure 1:**
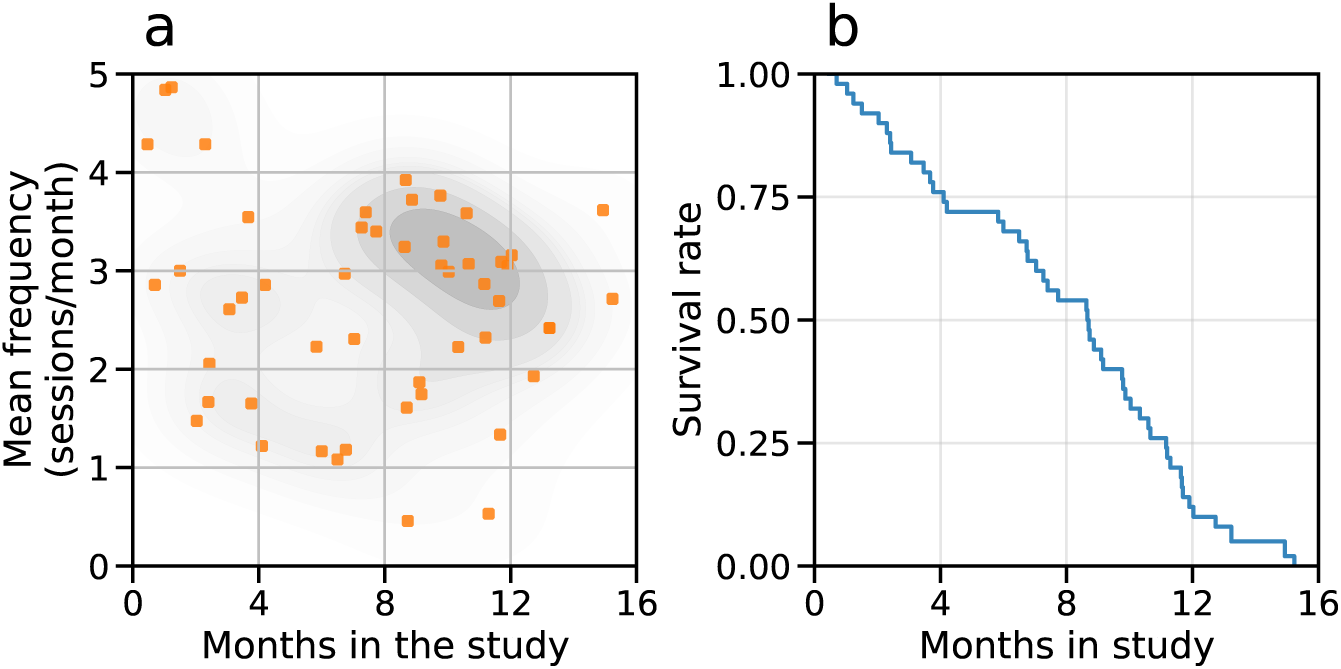
Participation in the Radcliff Study. (a) Distribution of participants based on their duration in the Radcliff Study and average session frequency. (b) Survival rate in the study, indicating that half of the participants were involved for 8.6 months or more.

Sessions were presented to participants weekly (see Methods). Ultimately, 55% of inter-session intervals ranged from 6-8 days and the average inter-session interval was 11.6 days (median 7 days). Maximum follow-up per protocol was 15.2 months. The mean follow-up duration amongst the entire population was 6.1 months (median 6.7 months) and in the subset included in this analysis, it was 7.7 ± 4.0 months (Figure 1b). The mean number of sessions per participant was 15.8 (range: 1 to 55) and the mean number of sessions per participant in the subset analyzed here was 20.3 (range: 3 to 55).

All FVC and SVC values reported in this work are expressed as percentage predicted values (see Methods). Each FVC or SVC measurement is recorded from the highest value from three consecutive trials. The quality of each trial is also recorded as *usable* or *non-usable*. Usability is determined by an automatically computed criterion ensuring that the patient initiated the exhalation properly, comparing the FVC with the volume of air extrapolated from the start of forced expiration to time zero (see Methods for details). We applied straightforward quality control to the data, retaining only sessions with two usable, non-zero efforts. Using this protocol, 96% of the 929 SVC sessions and 88% of the 991 FVC sessions were retained. The resulting time traces are available in Figure S2.

Progression is essentially linear for each participant over the observed period. The linear model showed a significantly lower mean RMSE than a quadratic one for FVC (5.26 vs. 5.86; t=-3.26, p=0.002, Figure 2a). For SVC, no significant differences were found (t=-1.88, p=0.068), but the linear model had a smaller mean RMSE (4.48 vs. 4.70, Figure 2a). Additionally, the evolution of both respiratory tasks is highly similar, with strong Spearman correlation values on the slopes (*ρ*=0.75, p<0.001, Figure 2b) and intercepts (*ρ*=0.97, p<0.001, Figure 2c) between FVC and SVC trajectories. This suggests that the potential difference between these two measurements may be only theoretical. In view of the similarity between the two pulmonary measurements, we focus on the FVC time series in the following analyses, while the results for the SVC series are available in the Supplementary Materials.

**Figure 2:**
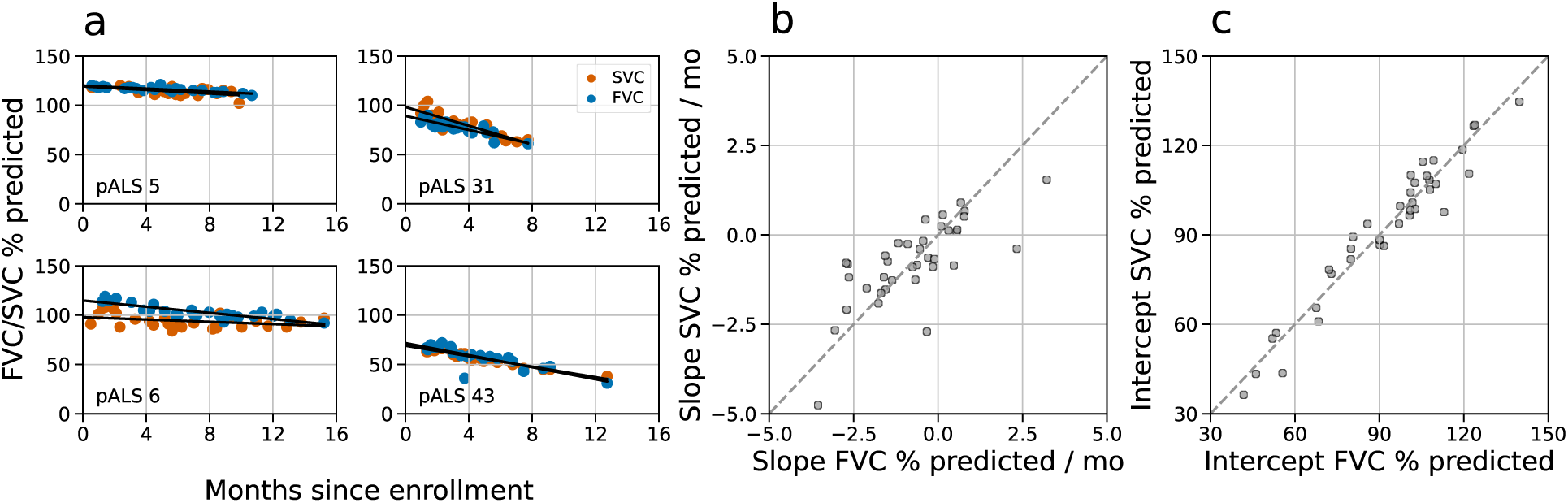
FVC and SVC time series evolve similarly. (a) Examples of the time evolution of FVC (blue) and SVC (brown) for pALS 5, 6, 31 and 43 (Figure S2), along with their linear regressions. Comparison of slopes (b) and intercepts (c) for the 37 pALS in the Radcliff Study with at least 6 FVC and 6 SVC sessions in the dataset and at least 2 months of valid sessions (Figure S1). High correlations for both the slopes (*ρ*=0.75, p<0.001) and intercepts (*ρ*=0.97, p<0.001) of FVC and SVC trajectories show consistency across time for both respiratory tasks.

The Radcliff study was designed for pALS to manage their coaching autonomously after an initial training period, allowing them to complete spirometry independently or request assistance from trained personnel (see Methods). This free-choice protocol yielded data of comparable quality for both proctored and non-proctored sessions: using the criterion of at least two usable, non-zero efforts per session, we discarded 12% of the 758 proctored sessions and the same percentage of the 233 non-proctored ones for FVC. Similarly, we discarded less than 4% of both the 703 proctored and the 226 non-proctored sessions for SVCs.

To compare respiratory progression between proctored and non-proctored sessions, we ran tests both at population and individual levels. At population level we performed a linear mixed model random slopes analysis for the proctored and non-proctored groups. No significant effect was found for the grouping variable (*p* > 0.9) or the interaction term (*p* > 0.8). At individual level, we calculated a linear regression for each pALS using all available sessions (proctored and non-proctored, Figure 3a). The root mean square error (RMSE) analysis revealed no statistically significant differences between the proctored and non-proctored sessions (*t* = −0.4, *p* > 0.69; Figure 3b). Additionally, we compared independent linear regressions for each pALS, one using only the proctored sessions and another using only the non-proctored sessions (Figure S3). We found positive Spearman correlation values for both the slopes (*ρ* = 0.85, *p* < 0.001, Figure S3b) and intercepts (*ρ* = 0.79, *p* = 0.002, Figure S3c), indicating good agreement between the progression of proctored vs non-proctored sessions. However, this comparison is limited to the cohort of 12 pALS with at least 4 proctored and 4 non-proctored sessions, over which slopes can be reliably estimated. Results consistent with these were obtained for SVC (see Figure S4 and Figure S5). This set of analyses builds confidence in the reliability and interchangeability of proctored and non-proctored respiratory measurements. Henceforth, we use both conditions altogether.

**Figure 3:**
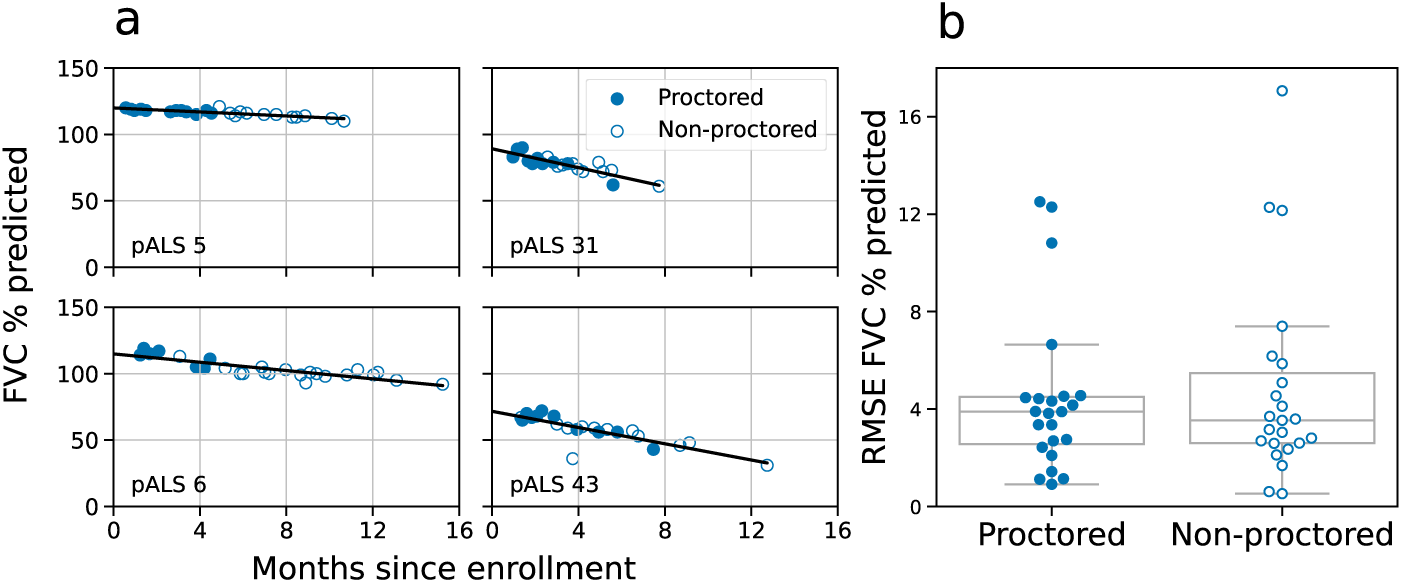
Proctored and non-proctored FVC spirometries. (a) Time evolution of FVC for pALS 5, 6, 31 and 43 (see Figure S2) with blue points representing proctored sessions and white points representing non-proctored. (b) Linear regressions were computed for each pALS with at least 3 FVC sessions, at least 1 proctored and 1 non-proctored in the dataset and at least 2 months of valid sessions (23 pALS, 457 sessions with 284 proctored and 173 non-proctored), showing no statistical difference in the RMSE between the two cohorts (T-statistic= −0.4, p>0.69, *N*=23).

To characterize progression types across our cohort, we used an unsupervised approach based on Gaussian processes to group patient trajectories into clusters (MoGP, see Methods). Four clusters were identified (see Methods), as shown in Figure 4. Following the classification reported in Amaral et al. (2024), we named these clusters with slow, moderate, and fast progressors. The blue and orange clusters include 18 slow progressors with slopes ranging from 0 to -0.4 points per month, while the green cluster comprises 12 moderate progressors with an FVC decline of approximately 0.9 points/month. Finally, the red cluster contains 4 fast progressors with an FVC decline of around 2.1 points/month (see Table 2).

**Figure 4:**
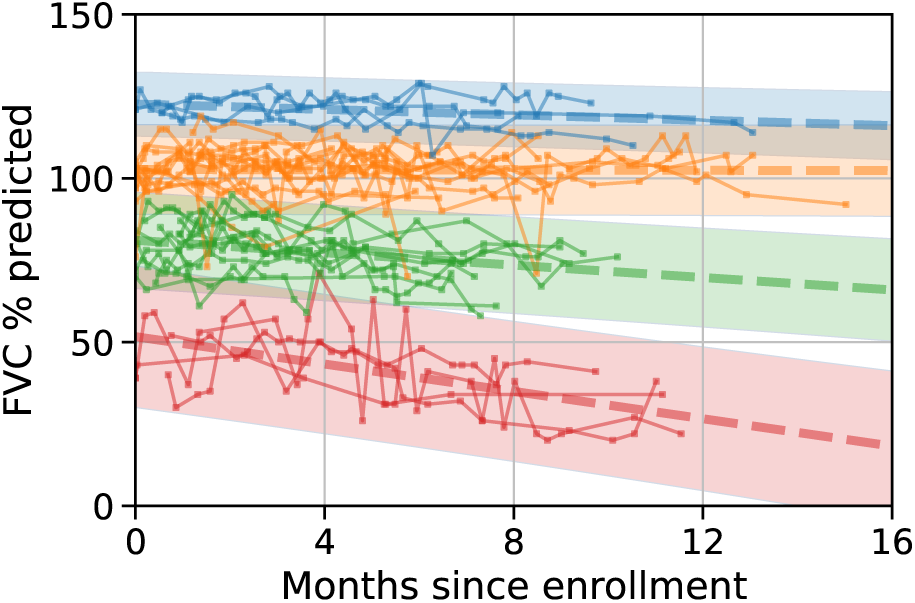
FVC progression types. (a) Similar trajectories are grouped using a MoGP linear model without predefining the number of clusters. The model identifies two clusters for slow progressors (blue and orange), for which FVC decreases less than 0.4 points/month, another one of moderate progressors (green) decreasing around 1 point per month, and one cluster of fast progressors (red), decreasing 2 points/month, see Table 2 for details.

**Table 2:**
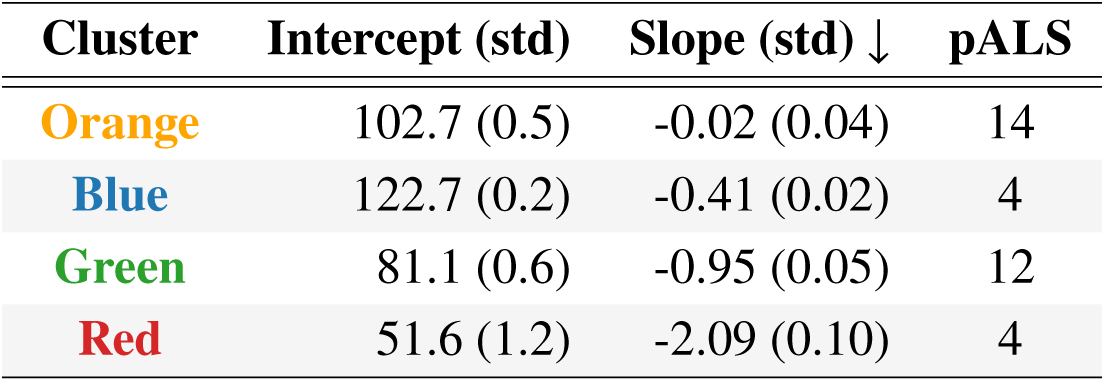
Parameters of MoGP linear clusters.

| Cluster | Intercept (std) | Slope (std) ↓ | pALS |
| --- | --- | --- | --- |
| Orange | 102.7 (0.5) | -0.02 (0.04) | 14 |
| Blue | 122.7 (0.2) | -0.41 (0.02) | 4 |
| Green | 81.1 (0.6) | -0.95 (0.05) | 12 |
| Red | 51.6 (1.2) | -2.09 (0.10) | 4 |

The identified clusters demonstrate alignment with the respiratory subscore ALSFRS-R (see Methods), reinforcing the validity of the proposed classification. Although no significant differences were observed for the total ALSFRS-R score (fast vs. slow: t=-1.436, p=0.114; moderate vs. slow: t=-0.255, p=0.401; fast vs. moderate: t=-1.284, p=0.134) or the Bulbar Subscore (fast vs. slow: t=-1.448, p=0.114; moderate vs. slow: t-statistic=-0.451, p=0.329; fast vs. moderate: t=-1.092, p=0.163), the respiratory subscore successfully differentiates between fast and slow progressors (t=-3.160, p=0.024) and between fast and moderate progressors (t=-2.492, p=0.032). In particular, the classification achieved by the MoGP model offers a higher level of granularity as it is capable of distinguishing slow from moderate progressors.

A relevant challenge in monitoring pALS is optimizing the duration and session frequency to accurately estimate disease progression. To address this, we compared the slope calculated over the entire study period with that estimated using only the first few months (Figure 5a). The difference between the ‘true’ and estimated slopes across participants is plotted in Figure 5b as a function of the number of months and session frequency. To explore the impact of session frequency, we resampled the data at 100%, 50% and 25% of the original sampling rate, corresponding to approximately weekly, bimonthly and monthly sessions, respectively (see Methods).

**Figure 5:**
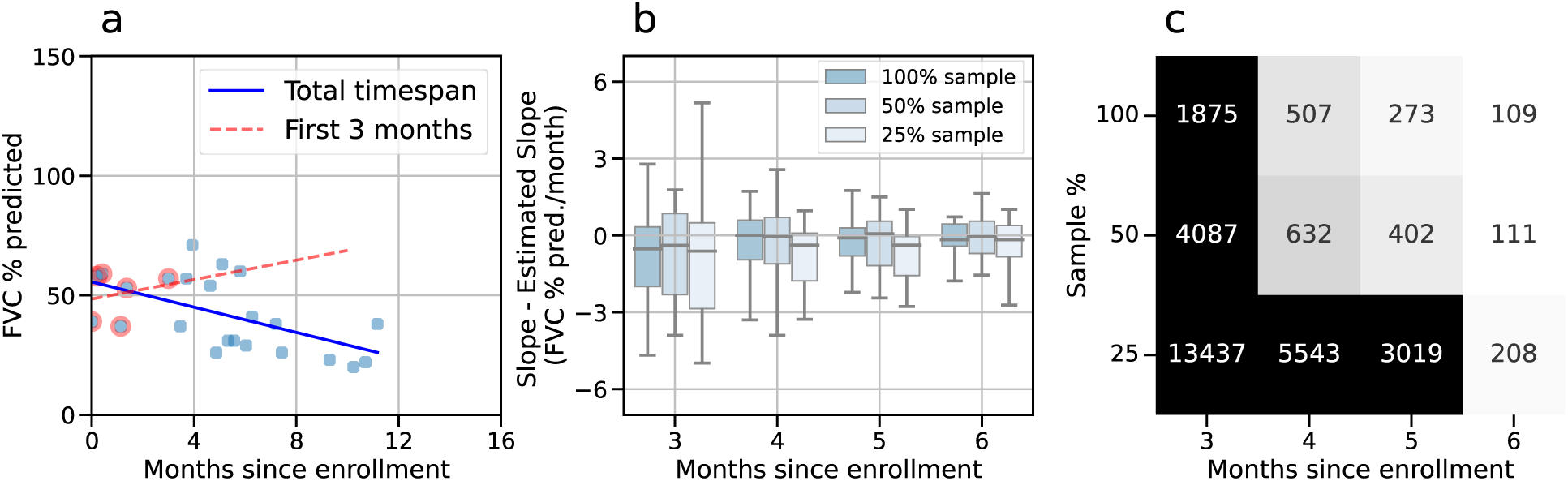
Predicting the FVC progression. (a) Linear regressions computed for FVC using the first three months of data (red), and the total timespan (blue). (b) The difference in slopes (%/mo) was calculated for the progressors (negative overall slope) as function of the number of months in the study. Results for all experimental data, mainly collected weekly, are shown in darker blue. Lighter tones represent the same results when the data are sub-sampled to bimonthly sessions (using one out of two sessions, 50% of the daata) and monthly sessions (using one out of four sessions, 25% of the data). (c) Estimated cohort sample size required for detecting a change of 30% in slope with 90% power at a significance level of 0.05, for weekly (100%), bimonthly (50%) and monthly (25%) sessions.

Interestingly, the difference between the ‘true’ and estimated slopes is often negative when only the initial months are analyzed. This results from a slight increase in FVC during the first sessions, which produces positive slope values—an effect that diminishes after the third month. This phenomenon can be multifactorial and may be associated with a learning effect; the fact that initial sessions are more likely to be proctored; or the disease takes longer to show decline.

The error decreases substantially for monitoring periods longer than three months and remains relatively stable across session frequencies (Figure 5b). However, while the error stabilizes across session frequencies after three months, the cohort size required to detect changes in slope varies considerably depending on duration and session frequency (Figure 5c). For example, increasing the study period from 5 to 6 months for weekly sessions reduces the required cohort size by nearly 60%, from 273 to 109 participants. In contrast, reducing the session frequency has a less pronounced impact. Halving the frequency to bimonthly sessions results in only a marginal increase in sample size (109 to 111 participants), whereas further halving to monthly sessions roughly doubles the required cohort size (111 to 208 participants).

## Discussion

In this study we demonstrate the feasibility and reliability of using home spirometry to monitor respiratory function in pALS, providing potential applications in both research and clinical practice.

Comparisons in the literature often highlight differences between remote and clinical spirometry (Anand et al., 2023; Paynter et al., 2022). As a fully remote study, the pulmonary tests performed in the Radcliff Study cannot be directly compared to clinical spirometry measurements from the same cohort. However, we present evidence that our flexible proctoring protocol, where participants performed respiratory tasks either independently or with remote guidance (Radcliff Study, 2024), generates at-home respiratory function measurements that are consistent across proctored and non-proctored conditions and comparable to clinical spirometry data from other studies.

First, we observed that SVC and FVC measurements are highly similar, aligning with recent clinical spirometry findings that demonstrate FVC and SVC decline are interchangeable for predicting functional decay in ALS (Pinto and de Carvalho, 2017, 2019). This similarity reinforces the robustness of both measures for tracking respiratory function, offering flexibility in their application depending on specific study or clinical needs. This is particularly relevant as FVC is generally used as the standard measure to evaluate respiratory function in ALS (Czaplinski et al., 2006), but some evidence suggests that SVC may be even more reliable (Huang et al., 2022).

We also show that both SVC and FVC decline linearly over observation periods of approximately eight months, supporting their use as reliable indicators of disease progression in ALS. This linearity is apparent not only at the cluster level but also in individual trajectories, providing an accurate basis for estimating progression rates during this time-frame. Notably, linear estimates of disease progression rates prior to enrollment in clinical trials serve as better predictors of patient survival than demographic data or discrete biological measures (Armon and Moses, 1998). While the long-term progression of pulmonary function in ALS is nonlinear (Panchabhai et al., 2019), the logistic patterns of FVC over extended periods (e.g., 80 months) can be approximated as linear within the shorter periods studied here.

Finally, the progression of pulmonary variables such as FVC has been studied. Although ALS progression is highly variable among individuals (Ramamoorthy et al., 2022), certain progression patterns of FVC have been identified in large longitudinal datasets (Ackrivo et al., 2019). Specifically, the study by Ackrivo et al. describes progression trajectories that are comparable in both baseline values and progression slopes to three of the clusters identified in our classification. Although we did not detect significant differences in the ALSFRS-R total score between the clusters, slow progressors had a higher ALSFRS-R respiratory subscore than fast progressors. The distinction between slow, moderate, and fast progressors based on FVC highlights the higher granularity provided by spirometry compared to ALSFRS-R. This underscores the importance of at-home monitoring for capturing detailed disease progression patterns.

These findings support the hypothesis that an adequate proctoring protocol, combined with an automated quality control process, produces at-home measurements of quality comparable to clinical spirometry. The ability to perform frequent monitoring through home spirometry offers significant advantages for clinical trials by enhancing statistical power while reducing the logistical and physical burden of in-clinic assessments. Our results indicate that weekly or monthly monitoring intervals during four months or more are sufficient to achieve reliable estimates of progression slopes. This reliability forms the foundation for the practical benefits of remote pulmonary function testing, such as reduced logistical burden and increased accessibility.

While home spirometry proves effective for monitoring respiratory function in pALS, there are also limitations that must be acknowledged. As the disease progresses, for instance, individuals may find it increasing difficult to independently perform spirometry tasks. In these cases, caregiver engagement will be necessary to ensure reliable data collection. Despite these challenges, this study underscores the value of flexible proctoring as a reliable tool for tracking respiratory function in ALS at home. Our findings contribute to the optimization of respiratory monitoring protocols, enabling improved accessibility, more frequent and faster testing, and ultimately enhancing care and outcomes for individuals with ALS.

## Methods

### Ethics Oversight

All aspects of the design and conduct of the Radcliff Study are done under the approval of the Western IRB. Every participant is presented with a digital written informed consent through a study portal and provides written documentation of informed consent prior to undergoing any study procedure. All participants consented to inclusion of their study data (for example, coded identifiers, demographics, ALS history and outcome measures, and speech samples) in a large speech database that can be accessed by researchers under appropriate data use agreements.

### Radcliff Study & proctoring

The Radcliff Study provides participants with two different, flexible engagement modules. After guided onboarding and verification of participants’ testing confidence within three proctored sessions, sessions can be performed autonomously with remote result monitoring by proctor or convened in weekly video sessions from the comfort of the participants’ homes under guidance of their permanently assigned proctor. Each session commences with the participant completing the ALSFRS-R, ARES and CPIB scores through the EverythingALS app. The proctor establishes baseline metrics for fatigue and shortness of breath using a self-assessment scale, while additionally inquiring about any changes or updates that might have occurred regarding medication, assistive devices and diagnosis.

Subsequently, participants are guided through the first respiratory task of three efforts of FVC and SVC. After completing the respiratory tasks, participants access the EverythingALS app to undertake a first, 8-10-minute-long speech task with weekly alternating scripts focusing on story recall, sustained phonation and verbal fluency trials. Following the speech tasks, participants perform three gait and balance assessments while utilizing ‘smart insoles’: a 25-foot walk, a 6-minute walk, and stair climbing to the point of fatigue. When returning to the continuously running zoom call, they engage in 5 to 7 minutes of a post exercise speech evaluation. The proctor then reassesses fatigue and shortness of breath, followed by a second set of three FVC efforts (up to six efforts to meet ATS acceptability), before concluding the session.

The proctor’s role is to train and remove technical obstacles while coaching remotely throughout each effort and assuring safety by monitoring symptoms like dizziness or imbalance, scheduling subsequent remote sessions and addressing any inquiry regarding the current session or the instantly available to participant test results (without providing medical advisory), while updating on milestones achieved within the study group as a motivational tool and context to quality of data and study goal.

### ALSFRS-R scale

All participants completed the Amyotrophic Lateral Sclerosis Functional Rating Scale-Revised scale (ALSFRS-R, a standard tool used to assess the functional status of individuals with ALS (Cedarbaum et al., 1999), evaluating bulbar, fine motor, gross motor, and respiratory functions through 12 questions, scored from 0 (no function) to 4 (normal function), with a total score of 48 for the complete scale and 12 for the bulbar and respiratory subscores.

### Reported FVC and SVC measurements

All FVC and SVC values reported here are expressed as a percentage of the predicted values for their age, height and sex, as described by Hankinson et al. (1999).

### Usable efforts

According to the spirometry standards (Graham et al., 2019) an FVC effort is defined as usable when the following conditions are met: 1) the volume of air that is back-extrapolated (BEV) from the start of the forced expiration to time zero must satisfy that BEV≤5% of FVC or 0.100 L, whichever is greater; 2) no evidence of a faulty zero-flow setting is detected, and 3) no glottic closure is detected in the first second of expiration.

### Linearity test

To assess the linearity of FVC and SVC trajectories, we applied two regression models—linear and quadratic—to predict FVC and SVC values, respectively, based on time since the first session. Leave-One-Out Cross-Validation (LOOCV) was performed for each subject to evaluate the root mean squared error (RMSE) of both models, and a paired t-test was used to compare their performance. We identified and excluded an outlier with an unusually high RMSE (pALS 28 in Figure S2). Inspection of this trajectory revealed a consistent increase in FVC across sessions, followed by an anomalously low FVC measurement in the final session, likely attributable to noise.

### MoGP clustering

We applied Mixtures of Gaussian Processes (MoGP) clustering to identify distinct progression trajectories within the cohort. The MoGP algorithm models each cluster as a mixture of Gaussian processes, allowing it to capture variations in both the mean trajectory and temporal dynamics between groups. Using this approach, we automatically segmented the cohort into clusters based on similarity in the longitudinal data patterns. This method captures progression trends, providing robust clustering for time series data with differing rates of change and noise levels across the cohort. The algorithm was applied to the 41 pALS with at least six valid FVC sessions, comprising a total of 809 sessions (Figure S1). The MoGP algorithm identified four clusters with at least three trajectories, which comprise a total of 34 pALS.

### Data Subsampling

We assessed the impact of varying sampling frequencies and observation time spans on FVC progression slope estimation. Sampling frequencies were set at 100% (all sessions), 50% (every other session), and 25% (one in four sessions), corresponding approximately to weekly, bi-monthly, and monthly sessions due to the original weekly schedule. Observation time spans ranged from 90 to 180 days in 30-day increments. For each frequency and time span combination, only sessions within the specified time interval were included, with sessions selected at regular intervals based on the sampling frequency. This resampling approach accounted for variability in protocol adherence.

### Sample size calculation

We calculated the sample size required to detect a 30% mean difference in slope for each possible time span and frequency considered, assuming a 90% power (1-*β*) and a 0.05 significance level *α*. Following the method described by Rutkove et al. (2020), we discarded patients with a positive slope in their complete trajectory, resulting in a population of 24 PALS (Figure S1). Given an estimation of average slope *s̄*, the standard deviation SE_*s̄*_, and the desired difference to detect, the sample size *N* was calculated as

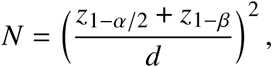

where *z* is the standard normal distribution for the respective level of significance and the power and *d* is the effect size computed as 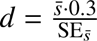.

## Data availability

All data needed to reproduce the figures in this work are available at the EverythingALS Reliable Monitoring RPFTALS Github repository.

## Acknowledgments

We would like to thank all the participants in this study who generously dedicated their time and effort.

## Supplementary Materials

**Figure S1:**
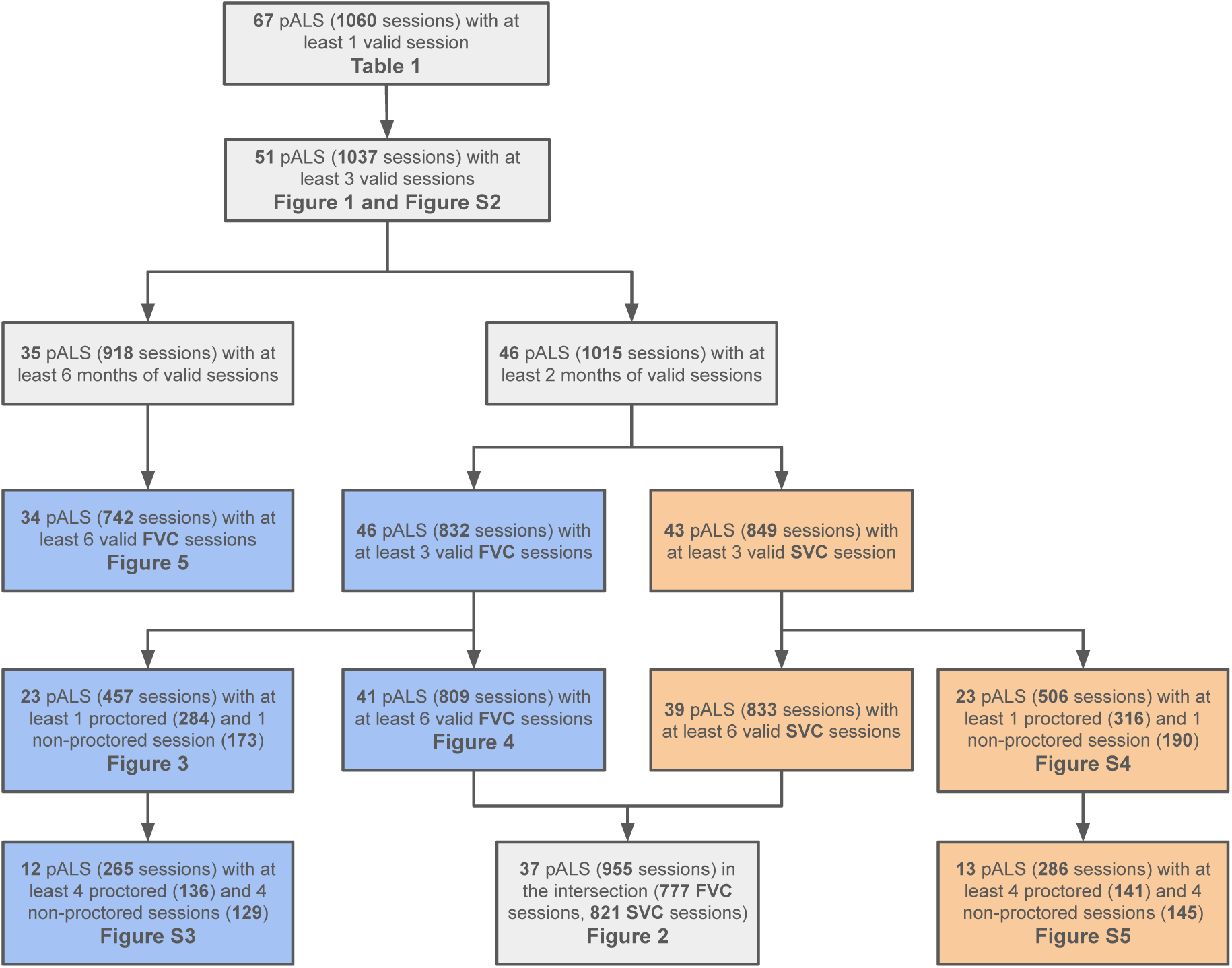
Overview of the data flow across the figures presented in this work. Blue and orange boxes correspond FVC and SVC data respectively.

**Figure S2:**
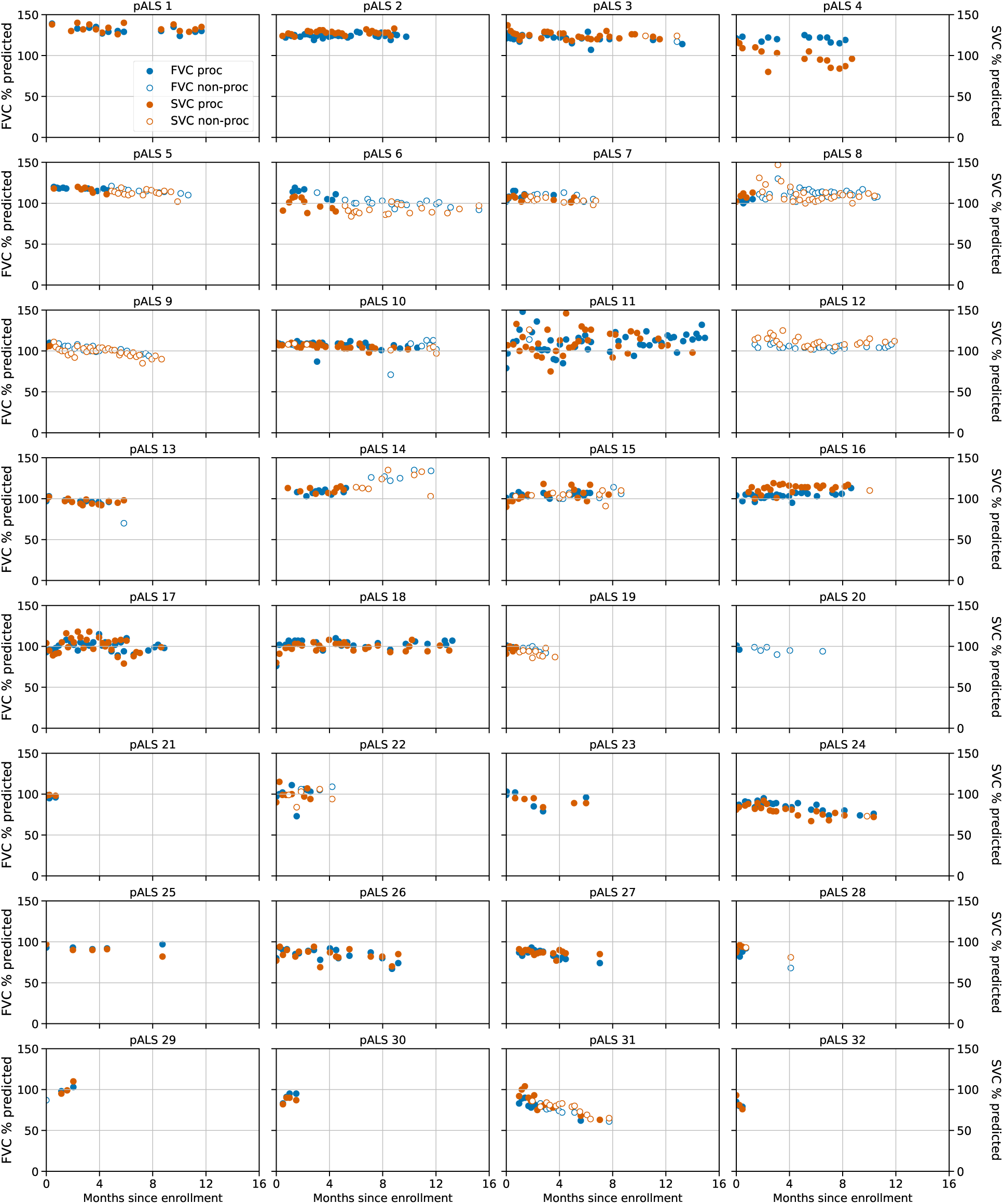
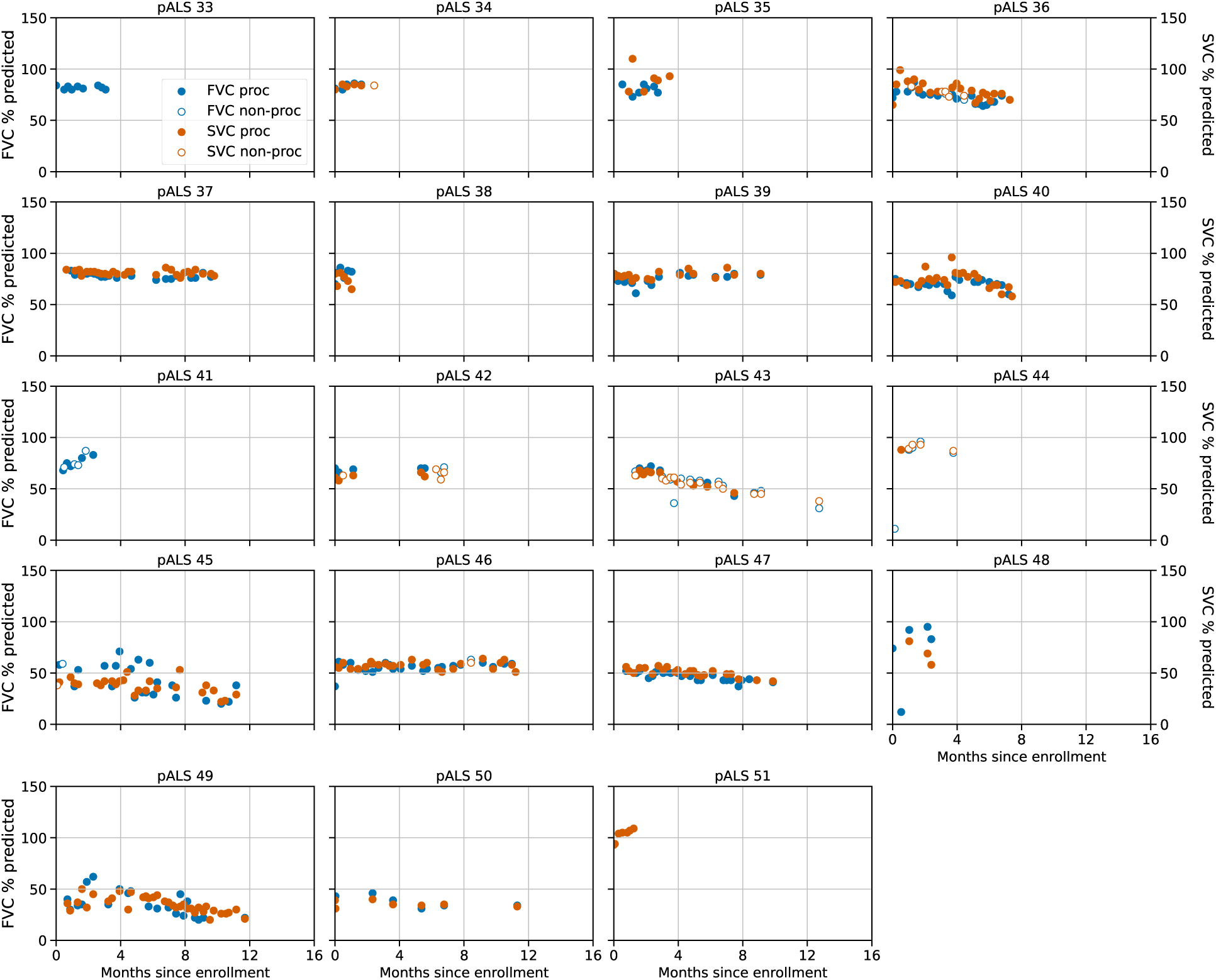
Evolution of SVC and FVC for pALS in the Radcliff Study. Trajectories of FVC are shown in blue and SVC in brown for the set of 51 pALS that completed at least 3 sessions (filled points are proctored, hollow non-proctored).

**Figure S3:**
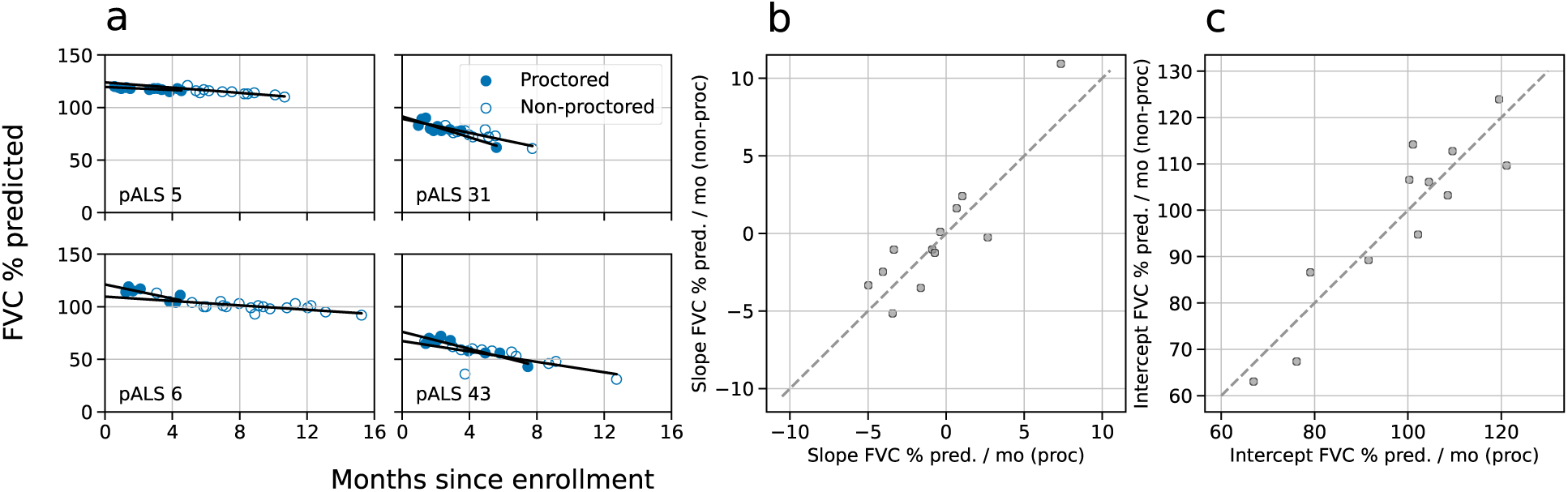
Individual regressions for proctored and non-proctored FVC data. (a) FVC time series examples for pALS 5, 6, 31 and 43 (See Figure S2) with linear regressions for proctored (blue) and non-proctored (pale blue) data after outlier removal. (b) Comparison of slopes and (c) intercepts for the 12 pALS in the Radcliff Study with at least 4 proctored and 4 non-proctored FVC sessions in the dataset (265 sessions). Good Spearman correlations for both slopes (*ρ*=0.853, p<0.001) and intercepts (*ρ*=0.797, p=0.002) show consistency across time for both conditions.

**Figure S4:**
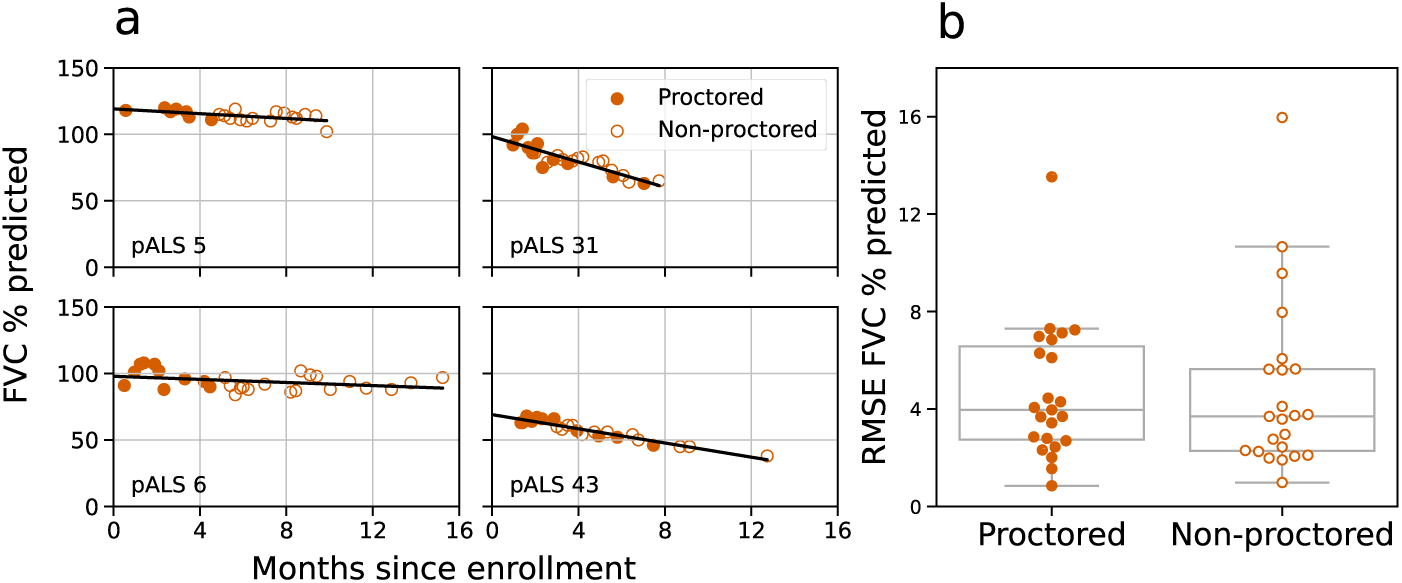
Consistency between proctored and non-proctored SVCs. (a) Time evolution of SVC for pALS 5, 6, 31 and 43 (see Figure S2) with orange points representing proctored sessions and white points representing non-proctored sessions. (b) Linear regressions were computed for each pALS with at least 3 SVC sessions, at least 1 proctored and 1 non-proctored in the dataset and at least 2 months of valid sessions (23 pALS, 506 sessions with 316 proctored and 190 non-proctored), showing no statistical difference in the RMSE between the two cohorts (T-statistic = −0.11, *p* > 0.9, *N* = 23).

**Figure S5:**
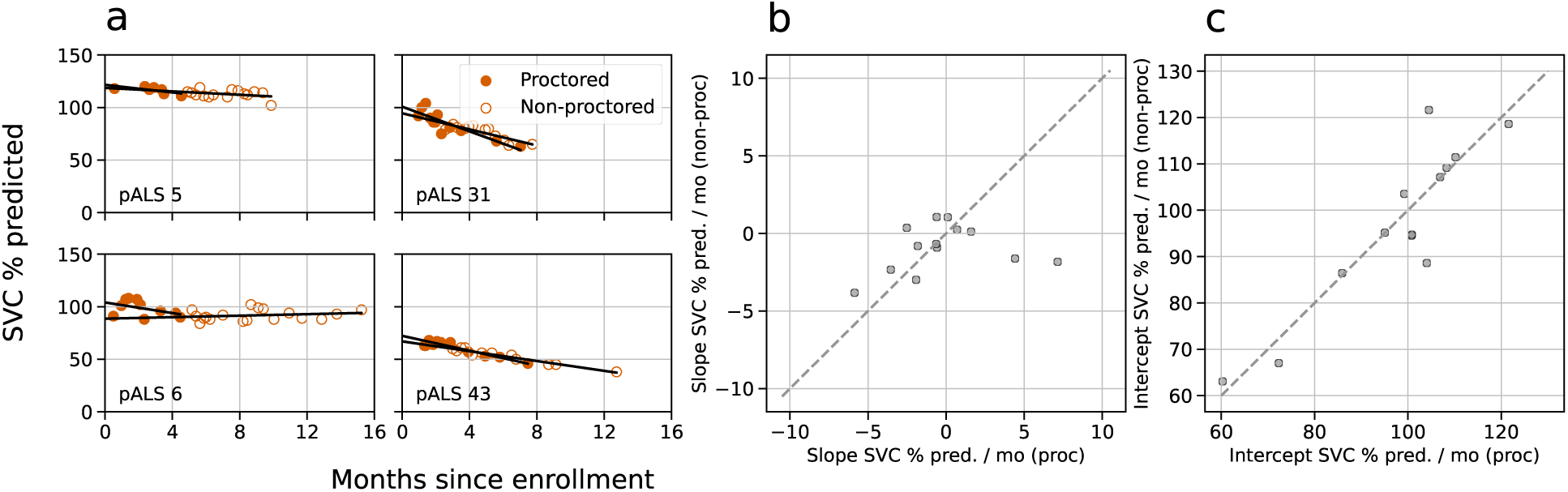
Individual regressions for proctored and non-proctored SVC data. (a) SVC time series examples for pALS 5, 6, 31 and 43 (see Figure S2) with linear regressions for proctored (dark) and non-proctored (light) data after outlier removal. (b) Comparison of slopes and (c) intercepts for the 13 pALS in the Radcliff Study with at least 4 proctored and 4 non-proctored SVC sessions in the dataset (286 sessions). Good Spearman correlations for both slopes (*ρ*=0.286, p=0.344) and intercepts (*ρ*=0.846, p<0.001) show consistency across time for both conditions.

## Notes

### Competing Interest Statement

JP, MAT, GB, FA and DES are paid consultants to EverythingALS. AT was a paid consultant to EverythingALS. Work was done while working at EverythingALS. Currently he is an employee of SRI International. SH and SB are paid consultants to EverythingALS. JO, RAS, PJ, JS are current employees of and report equity ownership in Bristol Myers Squibb. THP is a Medical Advisory Board for Mitsubishi Tanabe Pharma America, Amylyx and Novartis. She also received Clinical Trial funding from Novartis, and Clinical Study funding from Amylyx and Mitsubishi Tanabe Pharma America. MES declares no competing interests. MFW and OL are current employees and shareholders of Regeneron Pharmaceuticals, Inc. GS is an employee of Eli Lilly & Co. and holds company stock. KAS declares no competing interests. LWO and EF declare no competing interests. JDB has research support from Rapa Therapeutics, MT Pharma of America, ProJenX, Novartis, MDA, ALS Finding a Cure, ALS Association, Association for Frontotemporal Demenia and NINDS. He has been a consultant to Alexion Pharmaceuticals, Amylyx Pharmaceuticals, Biogen, MTPA, Regeneron Pharmaceuticals, Roon, Sanofi and Trace Neuroscience. He is an unpaid scientific advisor to EverythingALS. INB is the founder of EverythingALS. EGR is a paid consultant to EverythingALS.

### Clinical Protocols

https://www.everythingals.org/radcliffstudy

### Funding Statement

This study was funded by EverythingALS - Peter Cohen Foundation.

### Author Declarations

Salus IRB gave ethical approval for this work.

